# Strategies for Controlling the Spread of COVID-19

**DOI:** 10.1101/2020.06.24.20139014

**Authors:** Eric Forgoston, Michael A.S. Thorne

## Abstract

We consider a deterministic epidemiological compartmental model that includes age and social contact structure for the COVID-19 crisis and explore the consequences of different strategies for easing current lockdown measures that are in place in many countries. We apply the model to the specific circumstances in the state of New Jersey, in the United States of America. As expected, only a rigorous program of testing, tracing and isolation of cases will allow the state to ease its lockdown with a reduced number of deaths. We also find that a slightly earlier date of lockdown, while reducing the number of deaths in the short term, will only reduce the total number of deaths in the long run if the ensuing strategies in easing the lockdown are carried out with the aforementioned test, trace and isolation program. Otherwise, a slightly earlier lockdown will result in increased deaths as the expected second wave of infection sweeps through the state in the coming months.

## 1. Introduction

As the rapid spread of the coronavirus SARS-CoV-2 from its presumed origin of Wuhan, China has amply demonstrated, there has been a wide difference in the response to the pandemic from country to country. Many countries have responded rapidly with a degree of successful control, such as in South Korea [1] and Vietnam [2]. Other countries, notably the United Kingdom (UK) [3], the United States of America (USA) [4], and Brazil [5], have been particularly ill prepared to confront and deal with the emergency in timely fashion.

At the time of writing, mid June 2020, we enter the downturn of at least the first wave of the pandemic in many regions, with the centre of the pandemic slowly shifting to Africa, India and other parts of Latin America [6]. If adequate interventions are not put in place, the resulting casualty figures will most likely dwarf the current total. Meanwhile the fatigue of the crisis on the economy and individuals’ mental health is causing many governments around the world to push for a quick reopening and a return to business as usual. Unfortunately, this epidemic crisis is far from over, and short of a vaccine and a herculean vaccination program, we will be dealing with SARS-CoV-2 for a long while yet.

The standard way that epidemics are overcome is through widespread population immunity, otherwise known as herd immunity, preferably achieved with the aid of a vaccine. But without a vaccine, and an attendant large-scale program to vaccinate, the epidemic will run its course either through immunity of asymptomatic cases, recovered symptomatic cases and the death of critical symptomatic cases or, in the case of strong border and lockdown controls, eradication through quarantine, as recently achieved in New Zealand [7]. The latter is only effective if local or remote outbreaks are isolated from a disease-free population until these too are free of the virus through recovery or death.

Compartmental models are a standard deterministic epidemiological tool which have been used for over a hundred years to understand the outbreak and spread of infectious disease [8]. From a modelling perspective, considering the disease incubation time associated with COVID-19, a type of deterministic susceptible - exposed – infectious – recovered (*SEIR*) compartmental model is appropriate to investigate the dynamics [9]. Because asymptomatic transmission is so important in understanding the spread of SARS-CoV-2, we have extended the base *SEIR* model to include both asymptomatic and symptomatic individuals [10]. Since hospitalisation is playing a large role in the disease progression of COVID-19, this compartment has also been added [11]. We have chosen to include age structure in the model due to the wide discrepancy in hospitalisation and death rates between younger and older individuals [12]. Lastly, to understand the effect of implementing and easing different control strategies, we have incorporated social contact structure in the home, at work, at school, and in other places that are not home, work, or school.

We have chosen to parameterise the model using empirical data from New Jersey in the USA, and explore scenarios of implementing and easing social mitigation and lockdown strategies based on what has actually been done in New Jersey. While the specific outcomes of any given model is affected by the choice of parameter values and the control strategies used to reduce the ability of any contagion to spread, the progression of a disease follows clear patterns that are able to be transplanted across disparate circumstances. Even though we have considered as an application the situation in the state of New Jersey, the qualitative results are general and can be used to understand how the implementation and easing of lockdown measures will play out in other regions. In Section 2 we discuss the model and its parameterisation, while Section 3 considers the application of the model to New Jersey to better understand how the epidemic will evolve according to New Jersey’s choice of lockdown strategies as well as several possible scenarios of easing the control measures. Section 4 contains the conclusions.

## 2. The *SEI*_*s*_*I*_*a*_*HRD* model

Following the typical mathematical modelling of infectious disease using deterministic compartmental models, a susceptible - exposed - symptomatic infectious - asymptomatic infectious - hospitalised - recovered - deceased population model has been developed for individuals in 17 age groups given by [0 − 5], [5 − 10], [10 − 15], …, [70 − 75], [75 − 80], and [80+].

In this model, a schematic of which is shown in Figure 1, the population is divided into the following seven classes:

**Figure 1:**
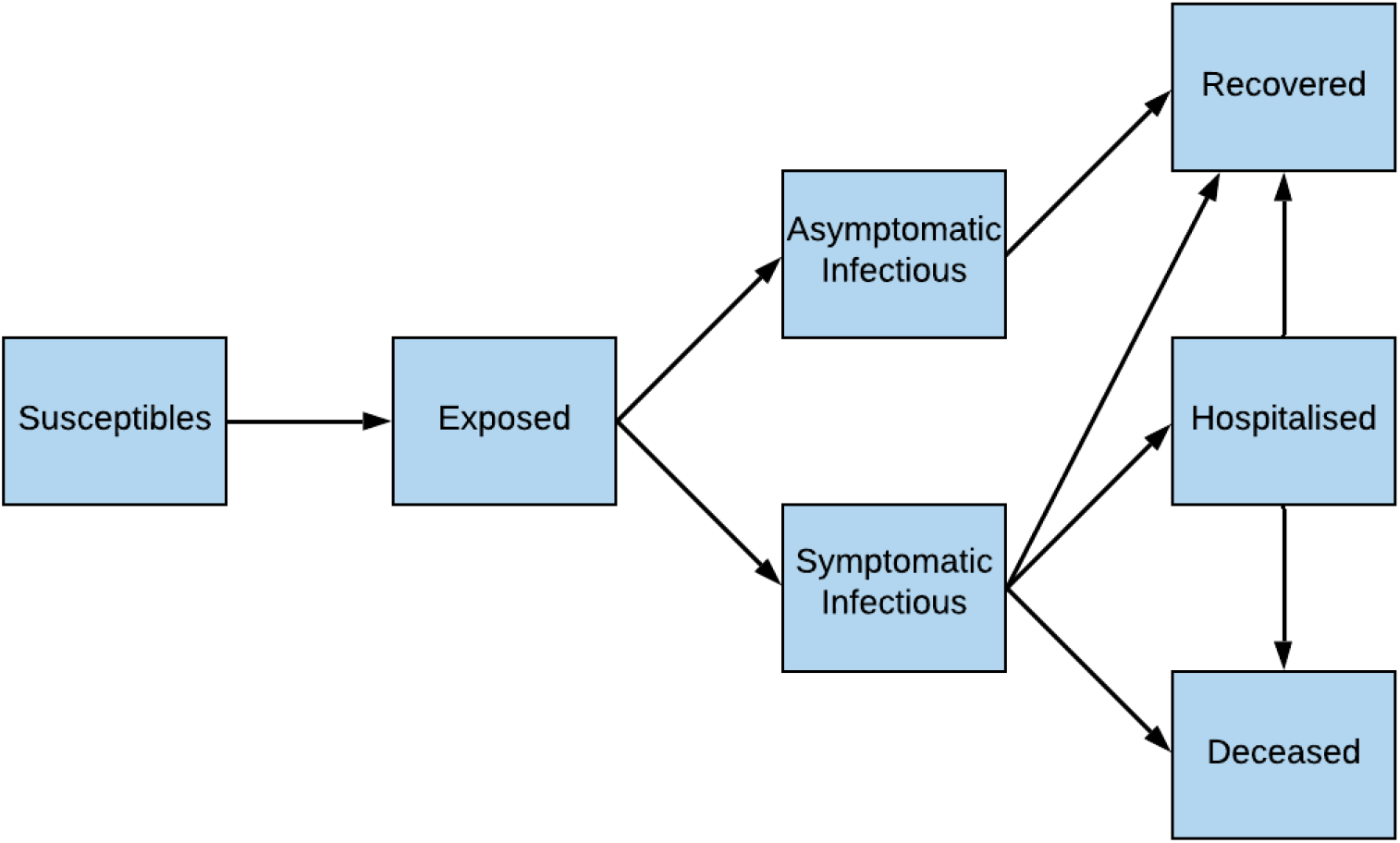
Flow diagram for the *SEI_s_I_a_HRD* COVID-19 compartment model.

1. *Susceptible* class *S* consists of individuals who may become infected with SARS-CoV-2 through contact with a symptomatic or asymptomatic infectious individual.
2. *Exposed* class *E* consists of individuals who are infected with SARS-CoV-2 but are not yet infectious.
3. *Symptomatic infectious* class *I*_*s*_ consists of individuals who develop symptoms of COVID-19 disease and are capable of transmitting the disease to a susceptible individual.
4. *Asymptomatic infectious* class *I*_*a*_ consists of individuals who do not develop symptoms of COVID-19 disease but nevertheless are capable of transmitting the disease to a susceptible individual.
5. *Hospitalised* class *H* consists of a fraction of symptomatic infectious individuals who must be hospitalised.
6. *Recovered* class *R* consists of individuals who have recovered from COVID-19.
7. *Deceased* class *D* consists of individuals who have died from COVID-19.

The governing equations for each of the *i* = 1 … 17 age groups are

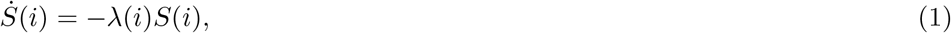

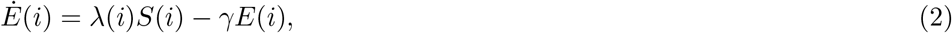

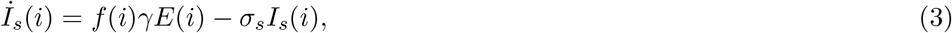

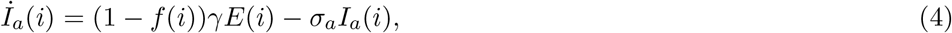

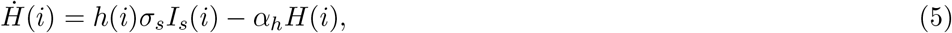

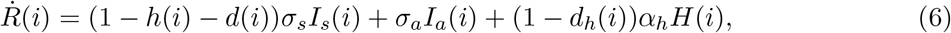

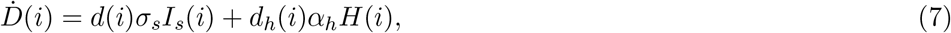

where

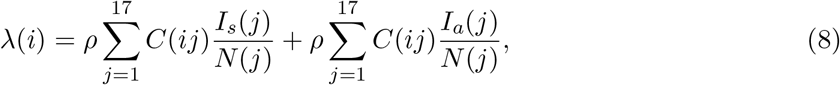

and where *S*(*i*), *E*(*i*), *I*_*s*_(*i*), *I*_*a*_(*i*), *H*(*i*), *R*(*i*), and *D*(*i*) respectively denote susceptible, exposed, symptomatic infectious, asymptomatic infectious, hospitalised, recovered, and deceased individuals for the *i*^th^ age group. In addition, *ρ* denotes the probability that a contact results in infection, *C*(*ij*) denotes the number of contacts of individuals in age group *j* with individuals in age group *i, N* (*i*) is the population size of New Jersey in each of the age groups, 1*/γ* is the mean exposure time, *f* (*i*) is the fraction of infected individuals who become symptomatic, 1*/σ*_*s*_ and 1*/σ*_*a*_ respectively represent the mean symptomatic and asymptomatic time, *h*(*i*) is the fraction of symptomatic infectious individuals who must be hospitalised, 1*/α*_*h*_ is the mean hospitalisation time, *d*(*i*) is the fraction of symptomatic infectious (non-hospitalised) individuals who die, and *d*_*h*_(*i*) is the fraction of hospitalised individuals who die. Parameter values that are specific to the physiology of COVID-19 and do not depend on age can be found in Table 1 while age-specific parameter values are given in Table 2. The social contact matrices for the USA are provided in [13]. The probability that a contact results in infection, *ρ*, was derived using empirically determined time-dependent values of New Jersey’s reproduction numbers [14] compared to the model reproduction numbers computed using the next generation matrix approach [15].

**Table 1:** Description of the various parameter values in the mathematical model which are specific to the physiology of COVID-19 and do not depend on age.

| Parameter | Description | Value |
| --- | --- | --- |
| $1/\gamma$ | mean exposure time (incubation period) | 5 days [18] |
| $1/\sigma_s$ | mean symptomatic time | 10 days [19] |
| $1/\sigma_a$ | mean asymptomatic time | 4 days [19] |
| $1/\alpha_h$ | mean (standard) hospitalisation time | 10.4 days [19] |

**Table 2:**
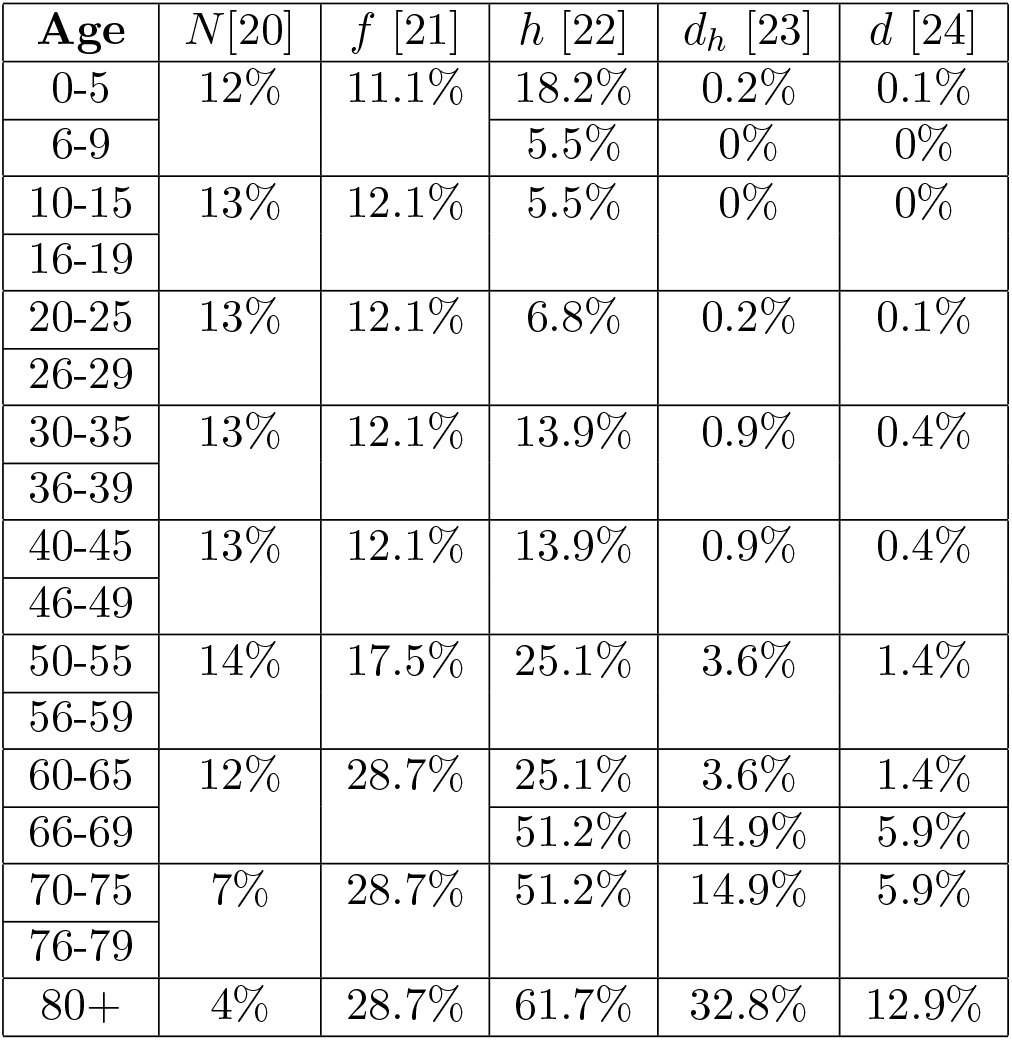
Description of the various age-specific parameter values in the mathematical model.

It is worth commenting that the model could easily be expanded to include a breakdown of types of hospitalisation, such as compartments for ICU outcome, with associated thresholds for numbers of ventilators or ICU beds after which fatality rates for critical hospitalised patients increase. However, the model needed to reflect the extent of available empirical data, and as we see in the next section, we have applied the approach to New Jersey, where the local statewide reporting did not provide the granularity to allow for a more detailed model.

It is also important to note that due to the current lack of adequate testing, in addition to the limited understanding of the physiological effects of the novel virus, there is great uncertainty in the parameter values, which leads to uncertainty in model simulations. In addition, we assume a number of facts about COVID-19 that remain unresolved, or for which the current research is still in dispute. Once recovered, we assume antibodies prevent reinfection. Also, we assume no mutating strain that alters the infection or fatality rates, despite some early evidence to the contrary [16]. Therefore, these rates remain constant throughout the epidemic’s progression. This work also predates the recent findings on dexamethasone, which by early indications will reduce death, if not hospital, rates substantially [17].

## 3. Application to New Jersey

Empirically-based contact information has been drawn on to determine the transmission of COVID-19 throughout New Jersey as well as the effect of lockdown measures and the removal or easing of different lockdown measures. The social contact matrices were obtained for the USA from surveys and Bayesian imputation [13] and are shown in Figure 2. The social contact matrices are divided into work, school, home, and other settings and show the average number of contacts between age groups in each of these settings.

**Figure 2:**
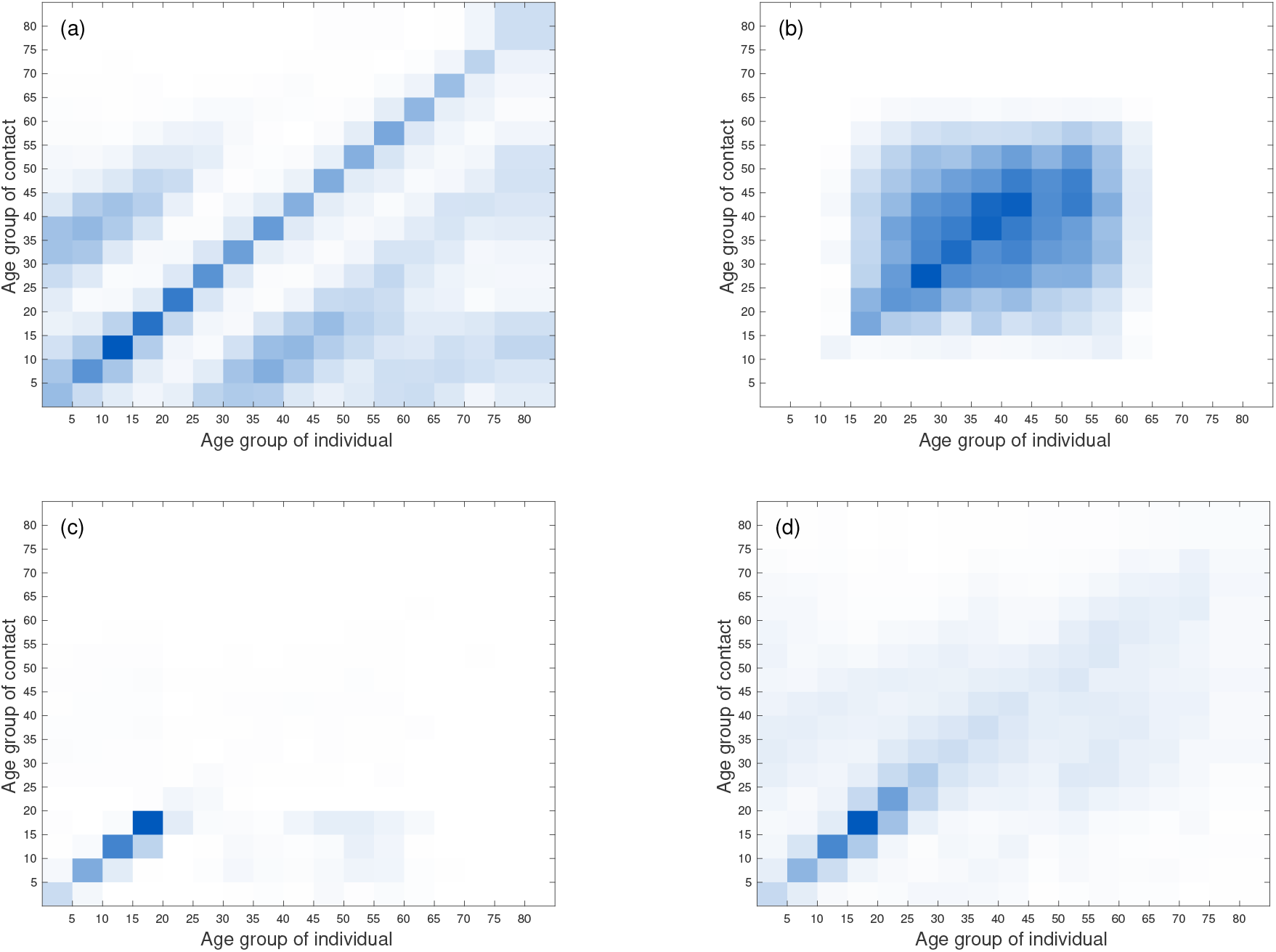
Age and contact structures of the United States of America at (a) home, (b) work, (c) school, and (d) other. The darker colours represent greater number of contacts.

By developing a COVID-19 model that includes age and social structure, we are able to assess the impact of specific social distancing and lockdown measures that have been implemented to contain the epidemic in New Jersey. To reflect the local (state-wide) spread of the pandemic, the model was parameterised for New Jersey by incorporating available empirical information. The available empirical data and parameter values for New Jersey are shown in Table 2 and were obtained from the New Jersey Department of Health [25], specifically the state’s information on COVID-19, as well as sites that have assessed the local reproduction number (i.e. [14]). When New Jersey specific data/values were unavailable, more general information from the wider COVID-19 literature and other comparable health services were used.

Figure 3 shows the number of symptomatic infectious individuals and cumulative deaths by age group for a model simulation that starts on March 04, 2020 with no lockdown measures. For comparison with outcomes due to various lockdown and easing of lockdown scenarios presented in Section 3.1, the inset figures show the total summed over all age groups. It is interesting to note in Figure 3(a) that because the oldest age groups are the smallest fractions of the total population (Table 2), these age groups also contribute the smallest numbers of infectious cases. However, the oldest age groups are disproportionately affected by COVID-19 with respect to hospitalisation and death rates, and Figure 3(b) shows the oldest age groups are responsible for the vast majority of the deaths.

**Figure 3:**
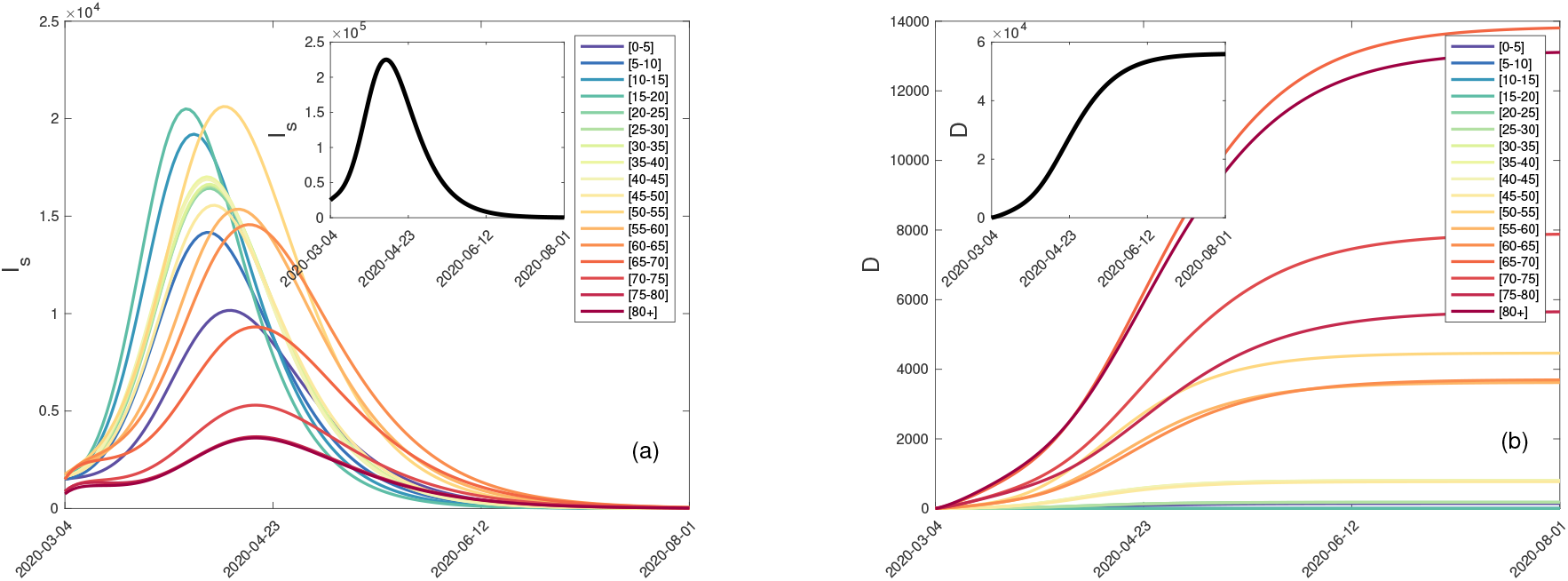
The (a) symptomatic infectious and (b) cumulative deaths for the *SEI_s_I_a_HRD* model without any interventions (the herd immunity scenario) applied to New Jersey broken down by the 17 age groups, with the total for each compartment inset.

### 3.1 Lockdown Easing Scenarios

The model enables one to assess the impact of removing specific lockdown measures on specific dates. Figures 4-6 show the number of infectious individuals, hospitalisations, and cumulative deaths. For each figure, the model simulations begin on March 04, 2020. Following New Jersey’s protocols, the model implements lockdown measures on March 16. In New Jersey as well as model simulations, the lockdown measures started to be incrementally eased on May 02 with the opening of state parks and golf courses. On May 18 construction resumed and curbside deliveries were allowed, beaches and lakeshores were reopened on May 22, and elective surgeries resumed on May 26. From this point, we explore different possible scenarios of removing the remaining lockdown measures (Figures 4 and 5), or re-implementing the lockdown measures when the number of infectious individuals increases (Figure 6). In all easing scenarios there is implicit in the resumption of normality an awareness of the need for social distancing either through the wearing of masks or separation of individuals. Therefore, the probability of disease transmission (determined empirically from NJ assessments of the reproduction number [14]) is lower than when no social distancing is occurring, as was the case before the pandemic began.

**Figure 4:**
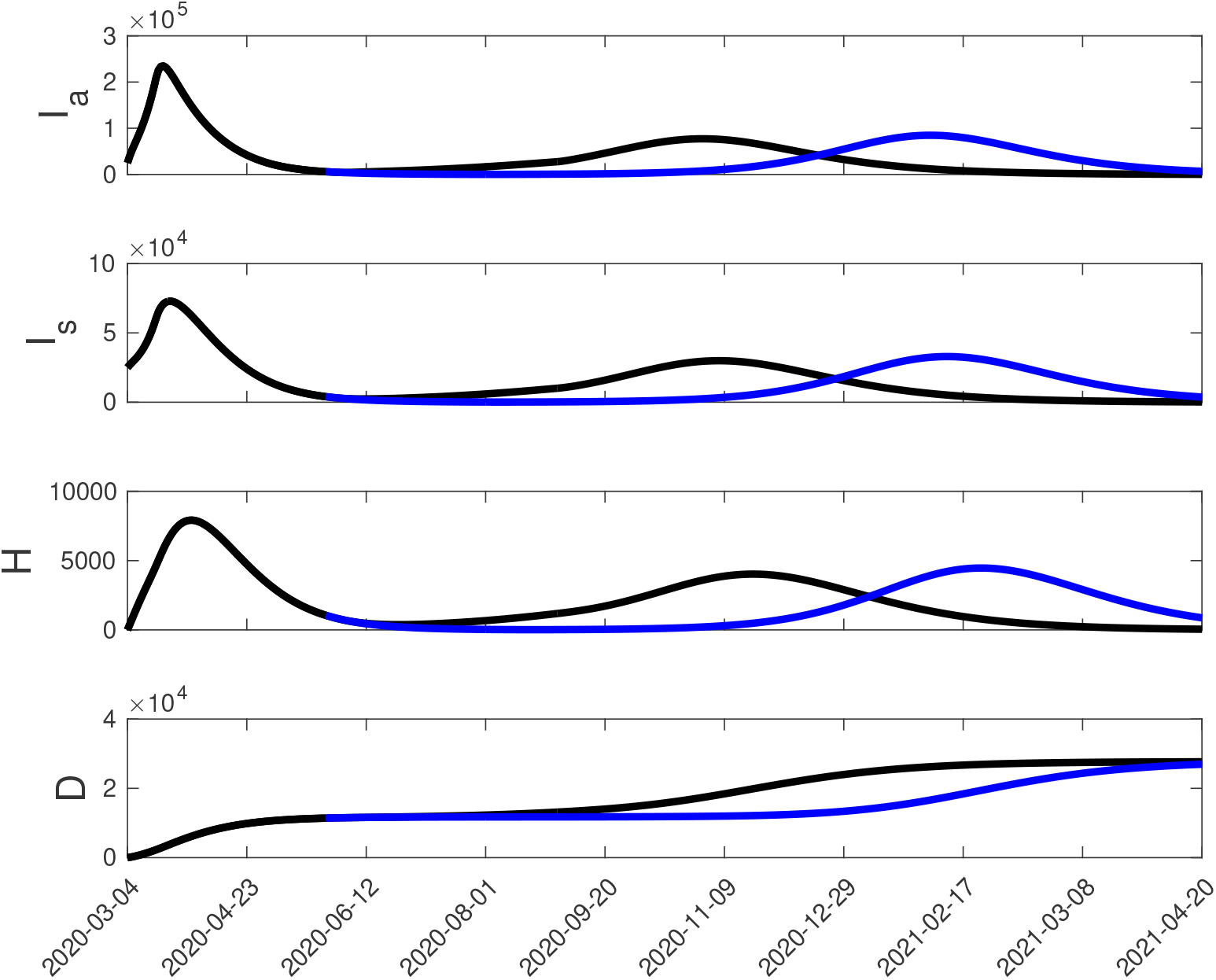
Number of asymptomatic infectious, symptomatic infectious, hospitalisations, and cumulative deaths over an approximately one year period beginning on March 04, 2020 with lockdown measures implemented on March 16. Starting on May 02 there began an incremental release of lockdown measures including the opening of state parks and golf courses on May 02, the resumption of construction and opening of curbside deliveries on May 18, the opening of beaches and lakeshores on May 22, and the resumption of elective surgeries on May 26. If all remaining lockdown measures are removed on June 01, with the exception of schools which are assumed to open on September 01 one sees the recurrence of a major second wave outbreak of infectious disease with a peak in November (black curve). If instead, the partially reopened state as of May 26 is maintained until August 01 at which point all remaining lockdown measures are removed including an early reopening of schools, one sees a similar second wave that is delayed so that the peak is in February, 2021 (blue curve).

**Figure 5:**
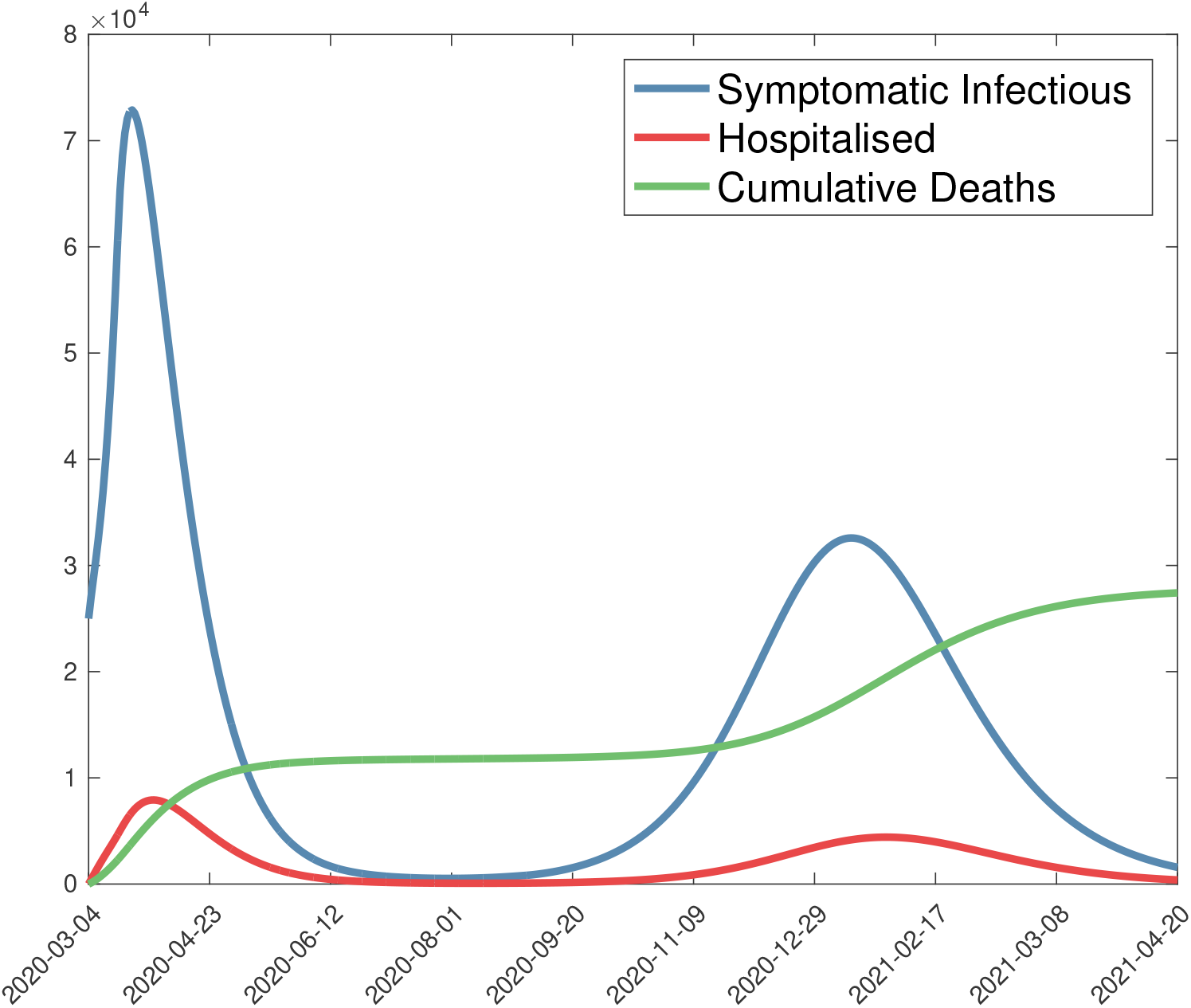
Number of symptomatic infectious individuals, hospitalisations, and cumulative deaths over an approximately one year period beginning on March 04, 2020 with lockdown measures implemented on March 16. Starting on May 02 there began an incremental release of lockdown measures including the opening of state parks and golf courses on May 02, the resumption of construction and opening of curbside deliveries on May 18, the opening of beaches and lakeshores on May 22, and the resumption of elective surgeries on May 26. If all remaining lockdown measures are incrementally removed every ten days throughout the summer until everything but schools are open by mid-August, and with schools assumed to open on September 01 one sees the recurrence of a major “second wave” outbreak of infectious disease with a peak in January, 2021.

**Figure 6:**
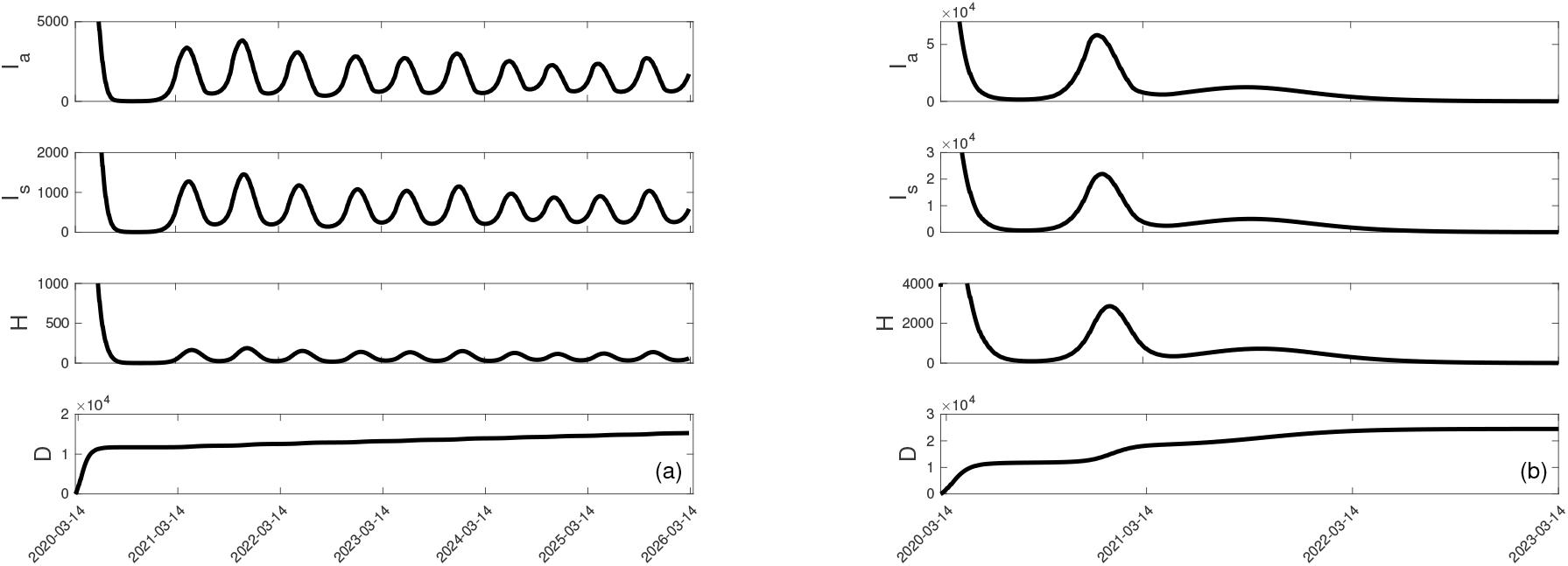
Number of asymptomatic infectious, symptomatic infectious, hospitalisations, and cumulative deaths over a multi-year period beginning on March 04, 2020 with lockdown measures implemented on March 16. Starting on May 02 there began an incremental release of lockdown measures including the opening of state parks and golf courses on May 02, the resumption of construction and opening of curbside deliveries on May 18, the opening of beaches and lakeshores on May 22, and the resumption of elective surgeries on May 26. After the slow relaxation of lockdown measures that took place in May, successive easing measures are continued at intervals of ten day periods. If the number of symptomatic cases increases above a threshold of (a) 10% more than the number of cases measured on May 22, or (b) twice the number of cases measured on May 22, then lockdown measures are successively tightened until the number of symptomatic cases falls below the threshold. At this time easing of lockdown measures resumes unless the number of symptomatic cases again rises above the threshold and tightening again begins.

Table 3 provides an overview of the number of fatalities depending on the form of easing of lockdown measures. If everything were to open on June 1, with schools opening in September, or if, instead, the lockdown continued until the beginning of August with early school openings, then the cumulative number of fatalities are approximately equal (Figure 4). A similar cumulative effect will occur with an incremental easing of lockdown measures throughout the summer (which is how NJ is reopening) with an opening of schools in September (Figure 5).

**Table 3:** Number of cumulative deaths for each model scenario. The number of fatalities as of May 22, 2020 determined by the model is 11322, whereas the empirical assessment as described by the NJ COVID-19 Daily Case Summary [26] is 11133. The cyclical easing/lockdown scenario is based on assessing the increase in new symptomatic cases relative to the number of symptomatic cases on May 22, 2020 and implementing increased lockdown measures in the event of an increase past the specified threshold, or conversely relaxing lockdown conditions if the number of symptomatic cases drops below the threshold. When considering cyclical easing/lockdown for thresholds larger than 700%, the implementation of lockdown or relaxation measures will result in the same number of cumulative deaths as that seen for the 700% threshold.

| Scenario | Cumulative deaths |
| --- | --- |
| **NJ Department of Health at May 22 [26]** | **11133** |
| Model at May 22 | 11322 |
| Open June 1 with schools opening in September (Fig. 4) | 27704 |
| Open August 1 with early opening of schools (Fig. 4) | 26895 |
| Successive easing of lockdown throughout summer (Fig. 5) | 27419 |
| Cyclical easing/lockdown based on threshold (10%) (Fig. 6) | 15289 |
| Cyclical easing/lockdown based on threshold (20%) | 18788 |
| Cyclical easing/lockdown based on threshold (50%) | 23294 |
| Cyclical easing/lockdown based on threshold (100%) | 24165 |
| Cyclical easing/lockdown based on threshold (200%) (Fig. 6) | 24496 |
| Cyclical easing/lockdown based on threshold (500%) | 26448 |
| Cyclical easing/lockdown based on threshold (700%) | 31133 |

All of the presented scenarios of lockdown easing result in further infections in the coming months, with most resulting in a substantial secondary wave. Only through close monitoring and isolation of infectious outbreaks will fatalities be kept down and a second wave give way to isolated, less substantial outbreaks (Figure 6(a)).

### 3.2. Consequences of an Earlier Initial Lockdown Date

New Jersey went into effective lockdown on March 16, 2020. Much has been made of the timing of the lockdown date in different regions, with a widespread criticism that it was too late. The first COVID-19 diagnosis in New Jersey occurred on March 4. Table 4 shows the number of fatalities under four of the scenarios described in the previous section. In the short term, on the specific date of May 22, an earlier lockdown date would clearly reduce the fatalities. If lockdown had taken place just two days after the first diagnosis, when there was ample evidence from elsewhere, including New York City, what would be coming, there would be a significant reduction by May 22 of 59% of the fatalities.

**Table 4:** Number of deaths dependent on an earlier lockdown date by stated scenario

| Lockdown date | At May 22 | August 1 easing | Successive easing | 10% cyclical |
| --- | --- | --- | --- | --- |
| March 16 | 11322 (100%) | 26895 (100%) | 27419 (100%) | 15289 (100%) |
| March 15 | 10691 (94%) | 28163 (104%) | 28426 (103%) | 14651 (95%) |
| March 14 | 10101 (89%) | 29163 (108%) | 29289 (106%) | 14077 (92%) |
| March 13 | 9548 (84%) | 30001 (111%) | 30055 (109%) | 13279 (86%) |
| March 12 | 9034 (79%) | 30731 (114%) | 30745 (112%) | 12642 (82%) |
| March 11 | 8557 (75%) | 31380 (116%) | 31370 (114%) | 11988 (78%) |
| March 10 | 8115 (71%) | 31963 (118%) | 31938 (116%) | 11499 (75%) |
| March 9 | 7708 (68%) | 32486 (120%) | 32451 (118%) | 10285 (67%) |
| March 8 | 7335 (64%) | 32954 (122%) | 32913 (120%) | 10378 (67%) |
| March 7 | 6997 (61%) | 33369 (124%) | 33324 (121%) | 9383 (61%) |
| March 6 | 6695 (59%) | 33733 (125%) | 33685 (122%) | 9154 (59%) |

However, an early lockdown needs to be complemented by a responsible lifting of lockdown measures. If the measures are lifted too quickly, as can be seen with the August 1st opening, or the successive easing throughout the summer (see Figure 7), during the second wave the fatalities are actually worse than a later initial lockdown. This may seem counterintuitive but is a consequence of more susceptible individuals post-lockdown that are able to become infectious where no further lockdown measures prevent COVID-19 from spreading. These scenarios do not assume a second or even further lockdown period. The cyclical easing/lockdown model however, with strict monitoring to catch any rising infections, can keep the fatalities down, so an earlier lockdown date coupled with strict enforcement of quarantine on any localised outbreak, or in the extreme case, further statewide lockdowns, would be the best approach to reduce fatalities.

**Figure 7:**
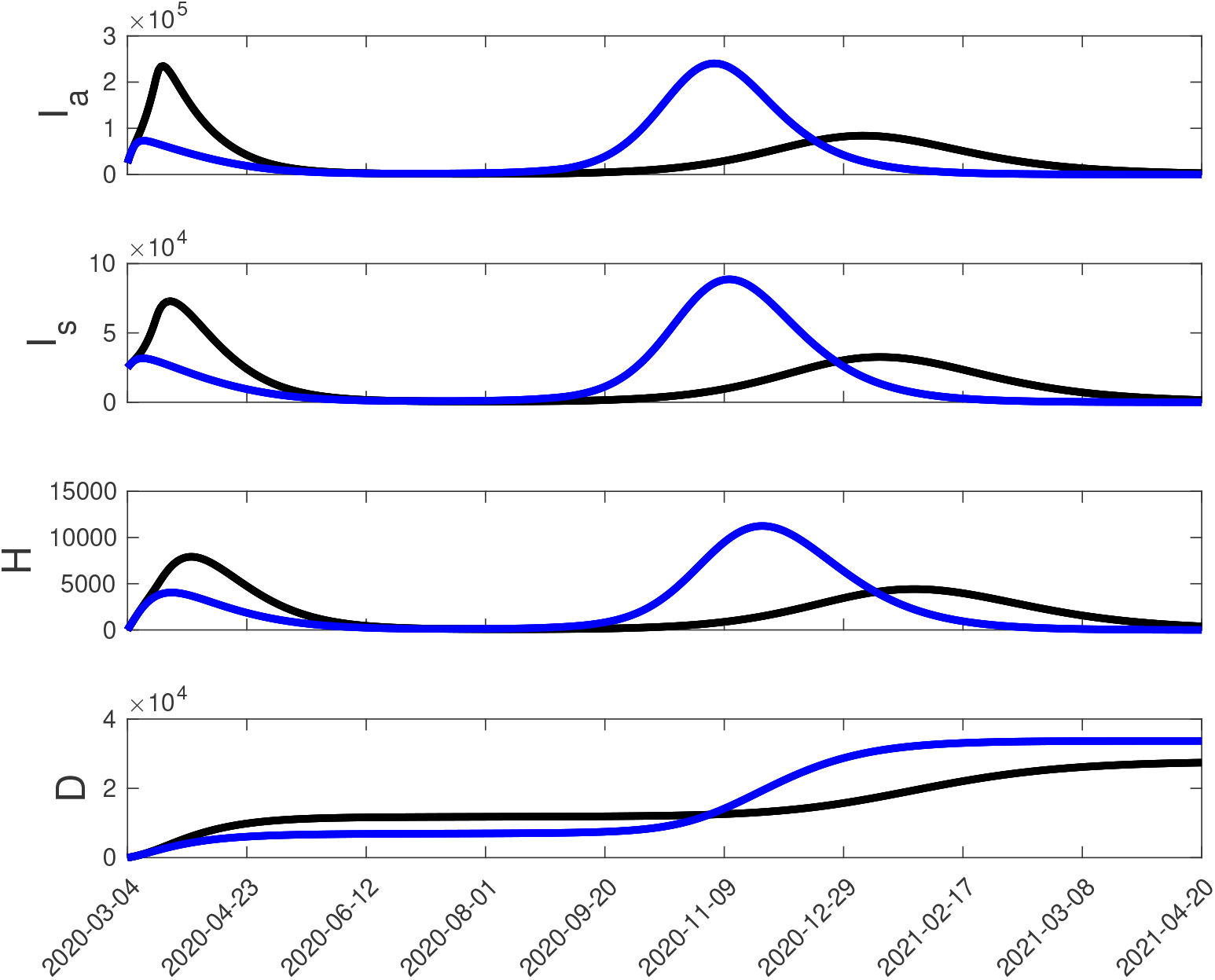
A consequence of an earlier lockdown. In black is the scenario shown in Figure 5, with the lockdown on March 16 and a successive easing of restrictions throughout the summer. In blue, we consider an earlier lockdown of March 6, two days after the first confirmed symptomatic case in the state. While initally a March 6 lockdown reduces the infectious rate, hospitalisations and deaths, as the lockdown is relaxed they actually increase in comparison to the March 16 lockdown date. This emphasises the importance of testing, tracing and isolating cases when easing lockdown.

## 4. Conclusion

We have developed a type of epidemiological compartment model useful for understanding the spread of COVID-19 in any region of the world. As an application, the model was parameterised based on empirical data from New Jersey, and was used to understand the immediate and long-term effects of different lockdown and easing strategies. The results demonstrate that to avoid a major second epidemic outreak, one needs to have adequate testing, contact tracing, and isolation/quarantine capabilities. The results also show that an earlier lockdown date will clearly have reduced the number of infections, and therefore also fatalities. However, any gains made by an earlier date can be lost by easing strategies that do not have in place by the time of relaxation a rigorous test, trace and isolation program.

In many regions, the reasons for lockdown was justified as a necessary measure so as not to tax the health services unduly, with concern for the number of ICU beds and ventilators and personal protective equipment and staffing among other considerations. While clearly this is an essential goal, and implicit in it is the saving of lives, the purpose of immediate lockdown when it was clear the pandemic was coming should have been to buy time. In other words, lockdown should have come very early, and while most citizens remain quarantined, the government and front line workers needed to scramble quickly to put into place a robust testing and tracing system so that on easing, any cases can be spotted and dealt with immediately, preventing further spread.

In South Korea, owing to previous experience with the 2003 SARS epidemic and the 2015 MERS epidemic, measures were put in place so rapidly that general lockdown was not even required [1]. With such an approach, coupled with other preventative measures such as hand washing stations, wearing of masks, and an information awareness campaign would have meant an earlier easing and much less disruption to the economy. The price for not having acted quickly and then continuing to ease without adequate measures in place means that there will be a second wave of infection. It would be hoped that lessons have been learnt during the first COVID-19 wave which can be employed to minimise the number of infections and associated deaths from further infectious waves.

## Data Availability

The code written to produce the models will be available upon request and through a github site.

